# Cortical Tau Aggregation Patterns associated with Apraxia in Alzheimer‘s Disease

**DOI:** 10.1101/2024.04.09.24305535

**Authors:** Gérard N. Bischof, Elena Jaeger, Kathrin Giehl, Frank Jessen, Oezguer A. Onur, Sid O’Bryant, Esra Kara, Peter H. Weiss, Alexander Drzezga

## Abstract

**Objectives:** Apraxia is a core feature of Alzheimer’s disease, but the pathomechanism of this characteristic symptom is not well understood. Here, we systematically investigated apraxia profiles in a well-defined group of patients with Alzheimer’s disease (AD; N=32) who additionally underwent PET imaging with the second-generation tau PET tracer [18F]PI-2620. We hypothesized that specific patterns of tau pathology might be related to apraxic deficits.

**Methods:** Patients (N=32) with a biomarker-confirmed diagnosis of Alzheimer’s disease were recruited in addition to a sample cognitively unimpaired controls (CU_1_; N=41). Both groups underwent in-depth neuropsychological assessment of apraxia (Dementia Apraxia Screening Test; DATE and the Cologne Apraxia Screening; KAS). In addition, static PET imaging with [18F]PI-2620 was performed to assess tau pathology in the AD patients. To specifically investigate the association of apraxia with regional tau-pathology, we compared the PET-data from this group with an independent sample of amyloid-negative cognitively intact participants (CU_2;_ N=54) by generation of z-score-deviation maps as well as voxel-based multiple regression analyses.

**Results:** We identified significant clusters of tau-aggregation in praxis-related regions (e.g., supramarginal gyrus, angular gyrus, temporal, parietal and occipital regions) that were associated with apraxia. These regions were similar between the two apraxia assessments. No correlations between tau-tracer uptake in primary motor cortical or subcortical brain regions and apraxia were observed.

**Conclusions:** These results suggest that tau deposition in specific cortical brain regions may induce local neuronal dysfunction leading to a dose-dependent functional decline in praxis performance.

## Introduction

Alzheimer’s disease (AD) is commonly strongly associated symptomatically with typical cognitive deficits such as memory loss, language and orientation problems and the loss of executive functions, whereas motor deficits are often less in the focus of clinical assessment. However, loss of higher order motor functions in particular is not uncommon in this disease. In particular, apraxia has been reported to be a core feature of AD, with a prevalence ranging from 35% in mild cases to 98% in severe cases (Lesourd et al., 2013). Generally, apraxia is defined as the “inability to perform specific and predefined actions or to execute learned and purposeful movements, independent of sensory, motor and (other) cognitive deficits” (Osiurak & Rossetti, 2017). Even in mild stages of the disease, the pattern of apraxic deficits can even help to distinguish the different variants of frontotemporal lobar degeneration and AD (Johnen et al., 2015, 2018). Apraxia can be considered to represent a highly relevant disability, as it can lead to impairment in activities of daily living and, thus, contribute to increasing care needs and loss of patient independence. Despite the high prevalence and clinical relevance of apraxia in AD, its pathomechanisms remain largely unresolved. Specific therapeutic options for apraxia remain generally sparse (Dovern et al., 2012). Current disease concepts of apraxia often rely heavily on evidence derived from studies on stroke. Apraxia is typically not considered to be a disorder of the core motor system which includes the primary motor cortex, basal ganglia, and cerebellum (i.e., not due to basic motor deficits such as paresis, ataxia, rigidity or tremor).

Instead, it is thought to result from a higher-order motor dysfunction of specific parieto-frontal praxis networks that constitute the praxis system (i.e., ventro-/dorso-dorsal and ventral action processing streams), or from a deficient interaction between the praxis networks and (praxis-related) cognitive networks (Goodale & Milner, 1992; Rizzolatti & Matelli, 2003). Apraxia is therefore often considered to be a higher-order motor-cognitive disorder. It is still unclear whether these concepts also apply to apraxia in AD, especially since there are very fundamental differences with the pathogenesis of stroke.

Apraxia in AD does not result from a on sudden neuronal injury, but rather from an insidious and progressive neurodegenerative process, leading to a gradual loss of function. In addition, AD pathology is typically bilateral and does not follow vascular territories. Importantly, AD neurodegeneration has often been thought to largely spare cerebral motor regions. Finally, the underlying neuropathologies are not comparable (i.e., ischemic injury in stroke versus protein aggregation pathologies in AD, involving amyloid plaques and tau tangles leading to neurodegenerative damage). Therefore, it seems important to analyze the molecular basis of apraxia in AD, to improve understanding of the underlying pathomechanisms and to possibly to open a different perspective on the development of apraxia in general.

Previous studies in AD suggest that tau-pathology in particular impairs with neuronal function in the brain in a region-specific manner (Dronse et al., 2016), as hyperphoshorylated tau-proteins dissolve from the microtubuli, aggregate intraneuronally in the form of neurofibrils, and are thought to contribute to local neuronal dysfunction. Modern in-vivo molecular imaging techniques, using positron emission tomography (PET) can non-invasively detect tau-aggregations in the human brain (Bischof et al., 2016). These methods may therefore be able to establish a specific link between regional AD-specific neuropathology and the resulting symptomatic deficits. In a recent study, we have already shown increased tau deposition in higher motor regions of patients with biomarker-verified AD pathology (Bischof et al., 2024). Analyzing tau PET scans from the Alzheimer’s Disease Neuroimaging Initiative (ADNI) cohort, we investigated how tau pathology in cytoarchitectonically mapped regions of the motor network varies across the clinical spectrum of AD. We were able to demonstrate significant tau pathology in predominantly higher motor regions in AD (e.g., supplementary motor area, superior parietal lobe, angular gyrus and dorsal premotor cortex, i.e., regions that have often been neglected). Based on these findings, we hypothesized that tau pathology, in praxis-related brain motor networks may contribute to apraxic deficits in AD (Bischof et al., 2024). We also suggested that the spatial heterogeneity of AD-pathology may contribute to the variability of apraxia severity in AD.

To test these hypotheses, we aimed to systematically assess apraxia in a well-defined group of patients with AD and correlate the results with data derived from tau-PET imaging in these individuals. To this end, we recruited a group of biomarker-confirmed patients with AD, who had been thoroughly characterized by neuropsychological assessment including systematic apraxia profiling using the Dementia Apraxia Test (DATE; (Johnen et al., 2018)) and the Cologne Apraxia Screening (KAS; (Latarnik et al., 2022; Wirth et al., 2016)). To reduce heterogeneity in the data, we planned to focus on patients with classical limb apraxia (deficits in gesture, imitation, pantomime, and actual object use), excluding other forms such as speech or gait apraxia. We performed standardized tau-PET imaging experiments in this population. For this purpose, we used the second-generation tau-PET tracer [18]F-PI-2620, which does not show relevant non-specific binding in the basal ganglia (Brendel et al., 2020), and evaluated how tau patterns in AD is associated with apraxia deficits.

With these experiments, we hoped to increase our knowledge of the mechanisms underlying the apraxic deficits in AD at the molecular level, and thus contribute to a better pathophysiological understanding that will also allow more sound diagnostic classification of this severe dysfunction.

## Methods

### Participants

The total number of participants was N=127. Two samples of cognitively unimpaired (CU_1_ & CU_2_) individuals were used in analyses described below. The first CU sample underwent behavioral testing only and was used to compute mean and standard deviation z-scores for the two tests measuring praxis function (CU_1_; N=41). These individuals were age-and sex-matched to the sample of AD patients and were included in the study if they (i) did not report subjective memory decline, (ii) were free of any psychiatric or neurological disease, (iii) had an MMSE above 26, (iv) were not taking any medication affecting the central nervous system.

The second sample of CU participants (CU_2_; N=54) underwent static imaging with [18F]PI-2620 only and were taken from the Health & Aging Brain Study – Health Disparities (HABS-HD) study population. Participant were included if they (i) did not report subjective memory decline, (ii) were amyloid negative on 18F-Florbetaben based on visual read and (iii) and had a CDR of zero. This group was used as a CU control sample to generate mean and standard deviation images to compute z-deviation maps for each AD-patient (details *see* imaging data pre-processing).

AD-patients were referred from the Center for Memory Disorders (ZfG; ‘Zentrum für Gedächtnissstörungen’). Patients were recruited based on the following inclusion criteria: (i) above 50 years old, (ii) biomarker confirmed amyloid pathology (i.e., positive visual reading on PET or decreased CSF beta-amyloid 42) and tau-pathology (i.e., positive visual reading on PET or increased CSF phosphorylated tau) according to recently proposed guideline (Bischof, 2022; Jack et al., 2018), (iii) ability to provide informed consent, (iv) no medical radiation exposure of > 60 mSv in the last 10 years, (v) no evidence of any form of dementia other than Alzheimer’s disease (vi) no other neurological or psychiatric disorder potentially responsible for cognitive decline or motor deficits (e.g. Parkinson’s disease). All AD patients underwent static imaging with [18F]PI-2620 and measures of tau tracer uptake were quantified (details see below Positron emission tomography (PET)).

The study was approved by the ethics committee of the Medical Faculty at University of Cologne and by the Federal Office for Radiation Protection (BfS), Germany. Written informed consent was obtained from all subjects prior to the study. Patient characteristics for the three different samples (CU_1_, CU_2_, AD) are shown in Table 1.

**Table 1).** Demographic information for cognitively unimpaired individuals (CU; behavioral sample; Cologne) and independent sample of cognitively unimpaired individuals (CU; Imaging controls - HABS-HD) where [18F]PI-2620 PET Data were collected. Demographic information on CU participants and the AD sample.

|  | *CU<sub>1</sub> (N=41)* | *CU<sub>2</sub> (N=54)* | *AD (31)* |
| --- | --- | --- | --- |
|  | Behavioral Controls<br>(Cologne) | Imaging<br>Controls<br>(HABS-HD) | Cologne |
| *Age* | 67.75 (8.94) | 66.62 (7.99) | 69.90 (10.98) |
| *Sex* | 22/41 (53 % male) | 30/54 (56 % male) | 19/31 (51 % male) |
| *MMSE* | 28.57 (2.0) | NA | 23.81 (4.09) |
| *DATE* | 56.39 (3.1) | NA | 46.00 (9.4) |
| *KAS* | 78.56 (1.4) | NA | 73.97 (6.0) |
| *CDR* | NA | 0 | NA |

## Materials

### Neuropsychological Assessment

The patients’ overall cognitive status was assessed using the Mini-Mental State Examination (MMSE), a test that evaluates impairments in general cognitive functions including orientation, attention, working memory, language, and delayed recall (Folstein et al., 1975). Impairment stages was defined based on previously published cut-off values (Folstein et al., 2000). Specifically, MMSE score of 20-26 indicated mild and 10-19 points moderate general cognitive decline.

The Cologne Apraxia Screening (KAS) was performed to characterize apraxia deficits in several categories including imitation of gestures, pantomime of tool use, and gesture recognition (Latarnik et al., 2022; Wirth et al., 2016). The Dementia Apraxia Screen Test (DATE) is a screening tool specifically designed to assess apraxia in individuals with dementia (Johnen et al., 2018). Similarly, to the KAS, it assesses aspects of gestures imitation, pantomime and tool use and gesture recognition, as well as aspects of buccofacial emblems and word imitation. The AD sample and the CU_1_ sample underwent these tests.

### Positron Emission Tomography (PET)

PET scans for the AD cohort were performed at the Department of Nuclear Medicine, University Hospital Cologne, Germany, with a Siemens Biograph mCT Flow 128 Edge scanner (Siemens, Knoxville, TN). All PET scans were iteratively reconstructed using a 3-D OSEM algorithm of 4 iterations and 12 subsets, postsmoothed by a Gaussian filter of 5 mm FWHM.

Patients were scanned for 30 minutes in listmode, 45 minutes after intravenous injection of 185 MBq of [18F]PI-2620 (FOV: 500 mm; gap = 1.5 mm, slice thickness = 3 mm) following recommended procedures for tau PET imaging with [18F]PI-2620 (Mueller et al., 2020). The PET images were reconstructed on a 128 × 128 matrix (3.18 × 3.18, slice thickness = 3 mm).

PET scans of the cohort of cognitively unimpaired individuals (CU_2_; N=54) were acquired on Siemens Biograph Vision 450 wholelJbody PET/CT scanner (O’Bryant et al., 2021). All participants were injected with a 10.0 mCi (±10%) bolus of [18F]PI-2620 and were scanned 45lJ75 minutes post injection. The PET images were reconstructed in a 440 reconstruction matrix with a zoom = 2, which results in 0.825 mm pixel (slice thickness = 1.64557 mm).

### Imaging Data Pre-Processing

All PET data was pre-processed with Statistical Parametric Modeling 12 (SPM12, Wellcome Trust Centre for Neuroimaging, Institute of Neurology, University College London). All PET images were aligned to the anterior-posterior commissure, co-registered to their corresponding MRI image and spatially normalized to the tissue probability map (TPM) implemented in SPM12. To create standardized-uptake value ratios (SUVRs) individual images were normalized by the non-specific binding of the vermis (Bischof et al., 2024). Finally, all images were smoothed with an 8 mm FWHM Gaussian filter. Despite differences in scanner acquisition parameters and subsequent differences in image reconstruction, standardization of pre-processing steps similar to the ones used in the Alzheimer’s Disease Neuroimaging Initiative (ADNI) PET Core (Jagust et al., 2015) ensured comparability.

### Imaging Data Post-Processing

All SUVR images from the CU_2_ sample were averaged and both a standard deviation and mean image were generated using Imcalc in SPM12. Mean and standard-deviation images were used to calculate z-deviation maps for each individual AD SUVR scan to create patient-specific AD-related tau patterns (Bischof et al., 2016). These images were the submitted for further analysis.

### Data Analysis

Demographic variables were analyzed using analysis of variance (ANOVA) and independent samples t-tests for continuous data and Chi-Square (*X^2^*) tests for frequency variables. Significance was set at *p* <.05.

### Behavioral analysis

To dissociate age-related differences in praxis function from AD-specific apraxia, we calculated mean and standard deviation values in the group of age-matched CU individuals (CU_1_) and calculated z-scores for each patient using the following equation:

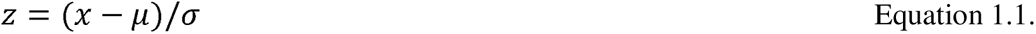

Resulting z-scores were entered as an independent variable in the multiple regression analysis described below.

### Imaging analysis

Mean deviation maps to display the typical pattern of AD tau pathology were created in SPM12. Individual z-maps from the AD sample were averaged using the Imcalc function (mean(X)). The resulting average map (*see* Results Figure 1a) were overlayed on the fsaverage image provided in the computational anatomy toolbox (CAT12) (http://dbm.neuro.unijena.de/cat/).

**Figure 1.**
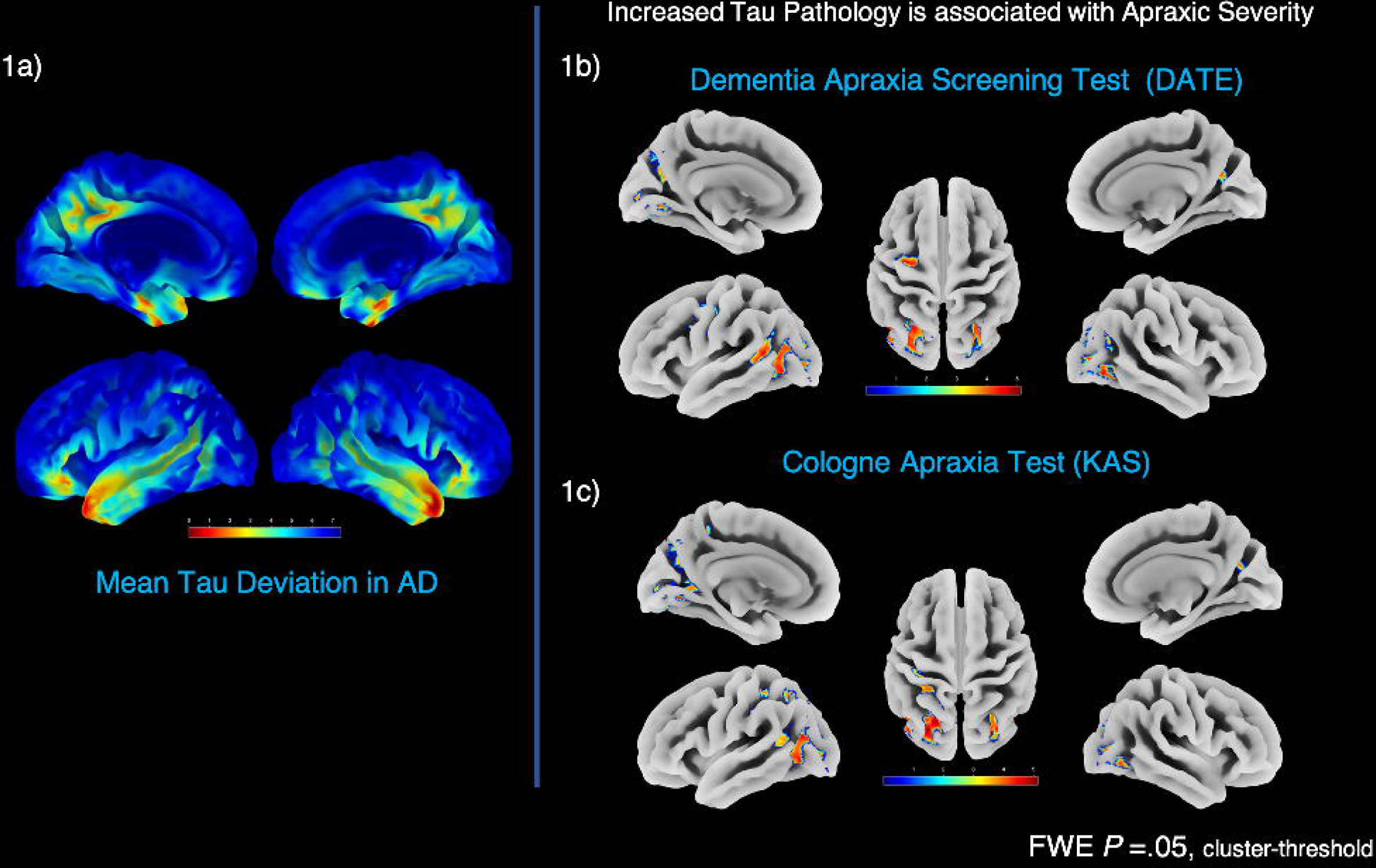
Caption: Results of the mean image of the deviation maps of the Alzheimer’s disease (AD) population (1a). Results from the two multiple regression models for the Dementia Apraxia Screening Test; DATE (1b) and the Cologne Apraxia Test (KAS) (1c). Family-wise error correction (FWE). Result maps are overlayed on a surface brain. Colo-Scale indicates z-values range in the voxels.

### Voxel-based regression analysis

For voxel-based analysis, individual z-deviation maps were subjected to multiple regression analysis in SPM12. Two separate models were constructed to fit the following equations:

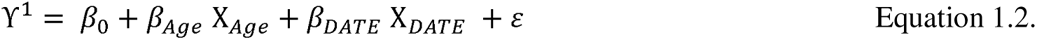

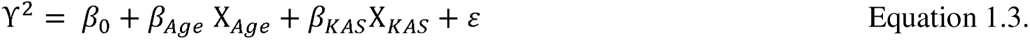

The statistical threshold was set at *p* = 0.05 with a family-wise error correction cluster threshold to account for multiple comparisons. The main contrast of interest was set to assess voxels of tau pathology associated with apraxia severity (DATE z-scores and KAS z-scores respectively), controlling for potential age differences. Results were overlaid on the Hammersmith Brain Atlas (Hammers et al., 2003) to quantify the percentage overlap of each significant cluster with anatomically derived regions.

## Results

Participant characteristics for the behavioral and imaging samples are shown in Table 1. One outlier from the AD cohort was removed, due to significant deviation (> 3SD) in both DATE and KAS scores. Adhering to the classification guidelines proposed by (Folstein et al., 2000), the majority of the sample of AD patients showed mild (N = 19; 61%) general cognitive decline, whereas a small proportion displayed moderate (N = 4; 2%) decline. Eight patients (N=8; 25 %) showed no impairment on global cognitive decline (MMSE > 26) despite biomarker positivity. Notably, none of the current patients was classified as having severe cognitive decline (MMSE < 10).

AD patients differed from the behavioral CU sample (CU_1_) in the MMSE and Apraxia scores (all *p* < .001), but groups did not differ significantly in age (t (69) = -.89, *p* = .37) or in the distribution of biological sex (*X^2^*(1, N=72) = 3.17, *p* = .07).

The range of deficits on the DATE test varied in the AD sample from 25 to 60, with 45 being the cut-off for significant motor impairment and 45 % of the sample scored below this cut-off. The KAS cut-off for apraxia is set at 76 and the range in the AD sample varied from 59 to 80, with 64 % of the sample showing apraxic deficits. Although some variation of motor impairment was found in the sample, the majority of people with Alzheimer’s disease had apraxia deficits on the KAS.

Both, the AD sample did not significantly differ from cognitively control samples (CU_1_ & CU_2_) in age (t (83) = -1.57, *p* = .12) or in the distribution of males and females within the group (*X^2^*(1, N=85) = 2.91, *p* = .08). Finally, age and sex also did not differ between the two control samples (i.e., CU_1_ & CU_2_) (t (92) = -.64, *p* = .52; *X^2^*(1, N=95) = 0.34, *p* = .85). Thus, both CU samples were sufficiently well matched to the AD sample.

Figure 1a shows the mean deviation map across all AD patients, which corresponds to the mean pattern of tau pathology that is distinct from the pattern of the group of cognitively unimpaired individuals (CU_2_). The spatial pattern of increased tau pathology included temporal regions, precuenus and cingulate cortex as well as inferior and superior parietal and rostral middle frontal and lateral orbitofrontal regions which is in correspondence to the spatial aggregation of tau in AD. No significant tau aggregation (i.e., z-score >2) was observed in basal ganglia in this analysis.

### Multiple Regression Analysis: Dementia Apraxia Test (DATE)

Several clusters of increased tau pathology were associated with apraxic deficits as assessed by the DATE. A surface overlay of the results map is shown in Figure 1b). The associated regions and their percentage overlap with the Hammersmith Brain Atlas are listed in Table 2a. Importantly, in addition to lateral occipital regions, apraxic impairment was associated with increased tau pathology in posterior and superior temporal regions, precentral and superior frontal regions, and the angular gyrus. Interestingly, these effects were observed in both hemispheres.

**Table 2:**
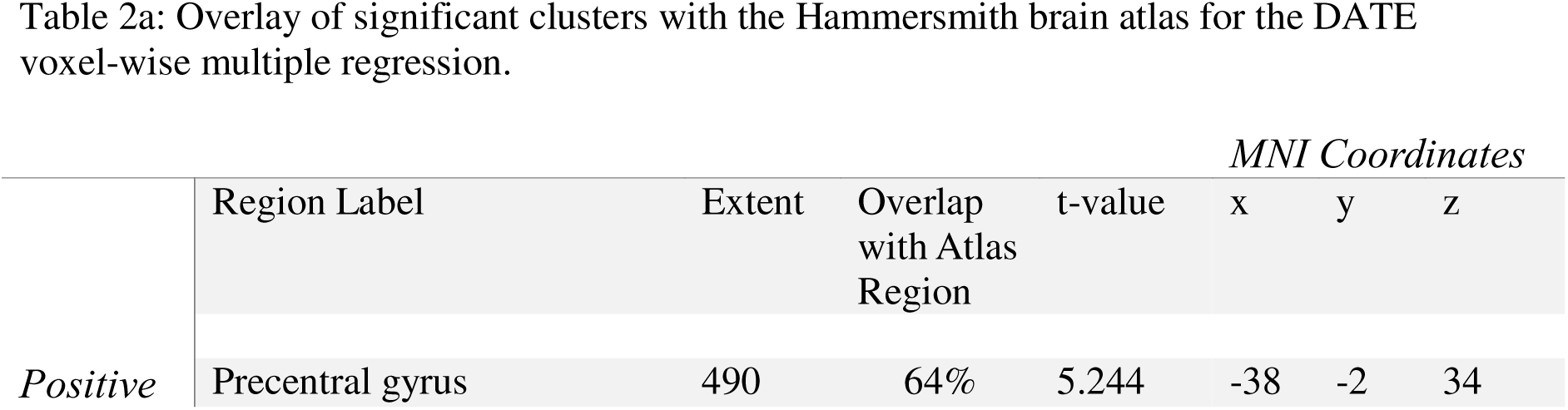
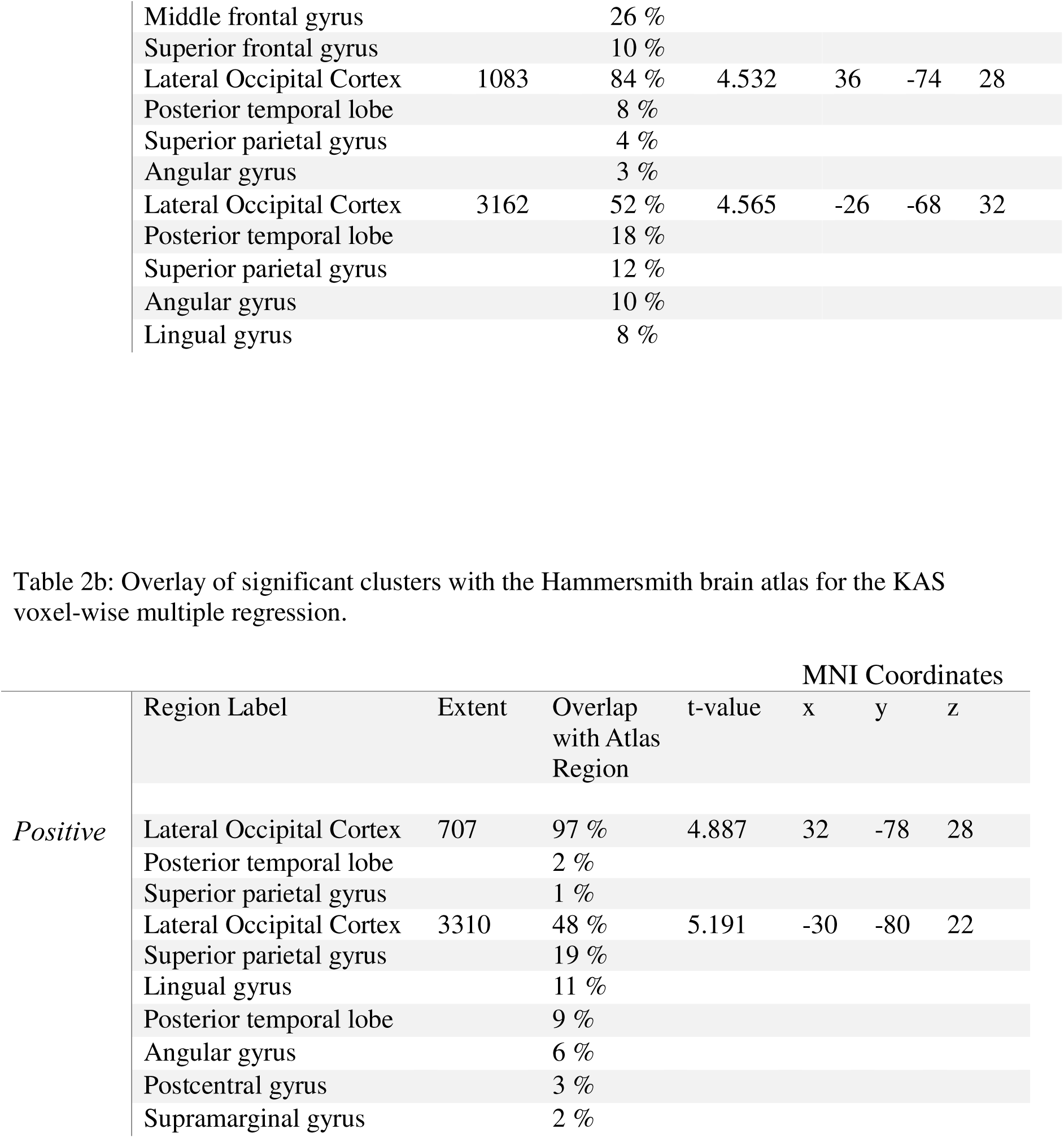

### Multiple Regression Analysis: Cologne Apraxia Screen Test (KAS)

A similar pattern emerged when assessing the regions where impairments on the KAS were associated with regions of tau pathology (see Figure 1c). Again, lateral occipital regions show a significant association with impairment on the KAS, in addition to superior and posterior temporal lobe, angular gyrus, supramarginal gyrus and superior parietal regions. The corresponding list of significant regions is shown in Table 2b.

Importantly, in both regression analyses, no correlation between tau-tracer uptake and apraxia symptoms was observed in primary motor cortical regions, nor in the basal ganglia.

## Discussion

In the current study, we were able to demonstrate for the first time that apraxic deficits in AD are associated with specific cortical tau deposition. Some affected brain regions (i.e. supramarginal gyrus, parietal regions, angular gyrus) are well-known for their involvement in praxis function (Martin et al., 2017b). This suggests that tau deposition in these brain regions may induce local neuronal dysfunction, leading to a dose-dependent functional decline in praxis performance. Importantly, we did not observe significant tau tracer retention in primary motor regions or the basal ganglia of patients, supporting the notion that apraxia in Alzheimer’s disease represents a higher-order motor dysfunction and is not caused by failure of the core primary motor system or subcortical involvement of the disease. We have recently demonstrated the involvement of tau pathology in motor regions in the context of AD severity (Bischof et al., 2024), whereas the current finding are extending our previous work by showing a direct association with apraxia symptomatology.

The group comparison of tau tracer uptake between AD and control subjects as quantified in the z-deviation map revealed a pattern of significant tau deposition in the brain, with maxima in brain regions including precuenus, posterior cingulate temporal pole, enthorinal cortex and inferior temporal regions, suggesting the group pattern of tau pathology extends beyond Braak III (Braak et al., 2011) . The pattern is consistent with previously described typical distribution patterns of tau pathology observed in AD (Bischof et al., 2016; Ossenkoppele et al., 2016; Schöll et al., 2016; Wang et al., 2016). These results serve as an indication that the included AD-population can be considered a representative sample and also that the employed control group used is suitable for statistical comparison.

Our findings are consistent with results from other studies, demonstrating high regional variance of tau-deposition patterns in AD and symptom-dependent association with tau-deposition in function-related brain regions (Bejanin et al., 2017; Brier et al., 2016; Vogel et al., 2021). First, an association has been shown between the overall severity of cognitive decline (as assessed by the Mini Mental Status Examination/MMSE) and tau burden, as measured in a large cortical meta-ROI as well as in the lateral temporal cortex (Bejanin et al., 2017; Boccalini et al., 2024; Brier et al., 2016). Importantly, pronounced tau-deposition in individual brain regions has been associated not only with overall cognitive impairment but also with specific decline of different cognitive functions associated with the respective brain areas (Dronse et al., 2016; Ossenkoppele et al., 2016). In particular, this refers to atypical variants of Alzheimer’s disease. These subtypes are defined by characteristic symptomatic features such as visual problems in the posterior cortical atrophy subtype, language problems in the logopenic aphasia subtype, or executive function problems in the frontal variant. Interestingly, the clinical symptomatic variability of these atypical forms of AD has been shown to be reflected in corresponding variation in the distribution of tau aggregation as measured by tau PET. Specifically, posterior/visual cortical regions were clearly affected in posterior cortical atrophy, asymmetric left hemispheric temporal accumulation was observed in logopenic aphasia, and pronounced frontal tau aggregation was observed in the frontal-executive variant, whereas bilateral tau tracer uptake in temporoparietal regions characterized typical AD (Dronse et al., 2016; Ossenkoppele et al., 2016; Phillips et al., 2018). Consequently, the authors of these publications concluded that the regional distribution of tau PET findings was strongly associated with the clinical and anatomical heterogeneity of Alzheimer’s disease.

Against this background, it seems plausible that the expression of apraxia, representing a motor symptom of Alzheimer’s disease, was found to be associated with tau aggregation in the corresponding function-relevant cortical brain regions. Tau-aggregation in regions belonging to the praxis networks showed a consistent correlation with apraxia severity as measured by two different rating scales (DATE and KAS), including the posterior temporal lobes, supramarginal gyrus, angular gyrus, and higher-order visual areas such as the lateral occipital cortex. The supramarginal cortex has been assigned to the ventro-dorsal processing stream/praxis network, which has been discussed to process action knowledge required for a proper tool/object manipulation (Kleineberg et al., 2018). The angular gyrus itself is considered to be relevant even for more than one praxis processing streams (Martin et al., 2017c). Additionally, posterior middle temporal gyrus/ middle occipital gyrus are part of the praxis network (Martin et al., 2017a) and do partially overlap with the clusters identified in the temporal and occipital regions here. Thus, a causal link between tau-induced neuronal dysfunction in these regions and observed apraxia symptoms appears in fact possible.

Importantly, the detected clusters do not overlap with brain regions where tau deposition has been associated with overall severity of cognitive decline, such as the lateral temporal cortex (Boccalini et al., 2024). Also, the parts of the praxis network involved are not among the brain regions affected by tau pathology early in the disease course (such as entorhinal cortex, Brodman area 35, neocortical temporal cortex (Berron et al., 2021)). It is therefore less likely that the associations found are simply due to more advanced tau deposition in the affected regions as the disease progresses.

To our knowledge, the severity of apraxia in Alzheimer’s disease has not previously been associated with specific regional tau pathology in Alzheimer’s disease. However, in a previous study by Palleis and colleagues, the tracer [18F]PI-2620 was used to distinguish subtypes of corticobasal syndrome (CBS) (Palleis et al., 2021). Higher tau levels, particularly in the dorsolateral prefrontal cortex, were observed in amyloid-positive CBS patients compared with an amyloid-negative group. Patients were also assessed for apraxia using the DATE. Interestingly, apraxia was found to be significantly more prominent in amyloid-positive than in amyloid-negative forms of CBS, supporting the notion that Alzheimer’s pathology is not infrequently associated with apraxia symptoms. In this population, however, no correlation between tracer retention and DATE apraxia scores was found. This may be due to the relatively small sample size of N=11 amyloid-positive subjects and/or lower symptom variation in this group of CBS. In this previous study, in both amyloid-positive and -negative CBS patients distinct tau-deposition in basal ganglia was observed (Palleis et al., 2021), which was not found in our current sample of AD patients.

Consistently, our study did not initially include patients with symptomatology consistent with CBS, but rather patients fulfilling criteria for typical AD (Jack et al., 2018). In particular, the difference in tau deposition in the basal ganglia between our sample and the group of CBS patients (studied with the same PET tracer) strongly supports the impression that AD with apraxia represents a distinct subtype and is not synonymous with amyloid-positive CBS. It can be considered a strength of the current study, that the employed tracer [18F]PI-2620 allows the assessment of tau-aggregation not only in the cortex but also in the basal ganglia, due to lacking non-specific uptake in these regions (Brendel et al., 2020).

This made it possible to demonstrate that motor deficits in AD can occur without the involvement of primary motor cortical (e.g., primary motor cortex) or subcortical (i.e., basal ganglia) brain regions. In our group of patients, apraxia was a surprisingly common finding, with abnormal scores detected in 64% of cases in the KAS and 45 % in the DATE. The severity of apraxia was not generally mild, but reached marked levels on the DATE and KAS, reflecting relevant disability. These findings underline that apraxia is a common symptom in Alzheimer’s disease that may require increased diagnostic attention and possibly therapeutic strategies. Cases with very dominant apraxia may even be considered to represent an "apraxic variant" of AD.

The current study has some limitations. First, the sample size of patients was still relatively small. However, all enrolled patients were well characterized and carefully matched to a set of controls. Nevertheless, replication of the results in larger samples of AD patients may be warranted.

We sampled different CU control groups to evaluate behavioral age-related apraxia scores and age-related tau pathology in amyloid negative individuals. Although these CU samples were matched by age-and sex to the target population, the different samples of CU individuals may have introduced a bias and our data may be less generalizable. The z-score deviation analyses approach however provided us with the same scale for both tau-pathology and apraxic score, which may reduce the influence on the different samples somewhat.

Finally, the CU_2_ sample, that provided the basis for the z-deviation analyses of the AD images, were scanned on a different scanner equipment compared to the AD cohort. Such differences can introduce a methodological bias which we tried to reduce by applying a smoothing kernel, and a standard matrix normalization, following pr-preprocessing steps from ADNI to reduce differences in scanners (Jagust et al., 2015).

In conclusion, to the best of our knowledge, this is the first paper to show a specific correlation of tau aggregation in praxis-related brain regions with the symptom of apraxia in a well characterized group of Alzheimer’s patients. The results suggest that tau in affected brain regions may contribute to local neuronal dysfunction and resulting functional deficits. Knowledge of these correlations could be taken into account in the diagnostic work-up and contribute to a better characterization and classification of patients with Alzheimer’s disease.

## Data Availability

All data produced in the present study are available upon reasonable request to the authors

## Acknowledgements

AD, GNB, EJ, MH and PHW are funded by the Deutsche Forschungsgemeinschaft - Project-ID 431549029 - SFB 1451. GNB received funding from Alzheimer Forschung Initiative e.V., Germany (AFI K1707). In addition, this study was supported by the German Research Foundation (DFG, DR 445/9-1). Precursor for the synthesis of the tracer [18F]PI2620 was kindly provided by Life Molecular imaging. The [18F]PI2620 control sample was kindly provided by Sid O Bryant and were taken from the HABS-HD database (https://apps.unthsc.edu/itr/studies/habs). Research reported on this publication was supported by the National Institute on Aging of the National Institutes of Health under Award Numbers R01AG054073, R01AG058533, P41EB015922 and U19AG078109. The content is solely the responsibility of the authors and does not necessarily represent the official views of the National Institutes of Health.

